# SCALE19: A scalable and cost-efficient method for testing Covid-19 based on hierarchical group testing

**DOI:** 10.1101/2020.06.01.20119172

**Authors:** Jeremie S. Kim, Justine S. Kim, Hyong S. Kim

## Abstract

Containment of Covid-19 requires an extensive testing of the affected population. Some propose global testing to effectively contain Covid-19. Current tests for Covid-19 are administered individually. These tests for Covid-19 are expensive and are limited due to the lack of resources and time. We propose a simple and efficient group testing method for Covid-19. We propose a group testing method where test subjects are grouped and tested. Depending on the result of the group test, subsequent sub groups are formed and tested recursively based on a quartery search algorithm. We designed and built an evaluation model that simulates test subject population, infected test subjects according to available Covid-19 statistics, and the group testing processes in SCALE19. We considered several population models including USA and the world. Our results show that we can significantly reduce the required number of tests up to 89% without sacrificing the accuracy of the individual test of the entire population. For USA, up to 280 million tests can be reduced from the total US population of 331 million and it would be equivalent saving of $28 billion assuming a cost of $100 per test. For the world, 6.96 billion tests can be reduced from the total population of 7.8 billion and it would be equivalent to saving $696 billion. We propose SCALE19 can significantly reduce the total required number of tests compared to individual tests of the entire population. We believe SCALE19 is efficient and simple to be deployed in containment of Covid-19.

## Background

Containment of Covid-19 requires an extensive testing of the affected population. Some propose global testing to effectively contain Covid-19. Current tests for Covid-19 are administered individually. These tests for Covid-19 are expensive and are limited due to the lack of resources and time. The concept of “group testing” was proposed in 1943 to efficiently detect syphilis among enlisted soldiers [1]. Group testing is also utilized in the field of bio-informatics and screening methods in biology [2]. Recently, the application of group testing is proposed for Covid-19 to reduce the number of tests [3,4]. The method described by Gossner et al. [3] tests a large group of people rather than an individual test. If the result of the test is negative, the entire group is deemed not infected. If the result is positive, then every member in the group is deemed infected. It does not identify the infected patient individually. Origami Assays [4] uses the combinatorial matching to infer infected patients from a large group of test subjects. It could potentially reduce the number of tests by 90% with a reasonable accuracy depending on the characteristic of group samples. However, for some cases, Origami Assays could have only 30% or less accuracy.

We present a group testing method that efficiently reduces the number of tests while also accurately detecting infected individuals from the testing groups.

## Methods

We propose a group testing method where test subjects are grouped and tested as a unit. For example, if a group consists of 16 patients, the required testing samples (i.e. blood, nasopharyngeal swabs) are collected from the 16 patients. These samples are combined into a “Group” sample and the COVID-19 test is administered to the “Group” sample. Depending on the result of the group test, subsequent sub groups are formed and tested recursively as follows. Assume there is a population of *N* test subjects. In this case, the required number of individual tests is *N*. Let *L* be the number of combined test samples within each group test. *N* test subjects are divided into multiple groups of *L* test subjects.

Now we describe our method for testing the group of *L* test subjects as shown in the flow chart in Figure 1. For simplicity, let *L* be 16. This group of 16 test subjects is tested. If the result is negative, then all subjects are deemed “not infected.” If the result is positive, then this group is divided into 4 groups with equal number of test subjects, in this case, 4 test subjects in each group. Then each sub group is tested and the process repeats until it reaches to an individual test subject. In this simple example, for 16 test subjects, only 9 tests are required as opposed to 16 individual tests. Thus, the total number of tests is reduced by 43%. Let *T_r_* be the required number tests to identify infected test subjects. As the probability of infection among test subjects gets lower, the cost saving increases significantly by reducing *T_r_*.

**Figure 1.**
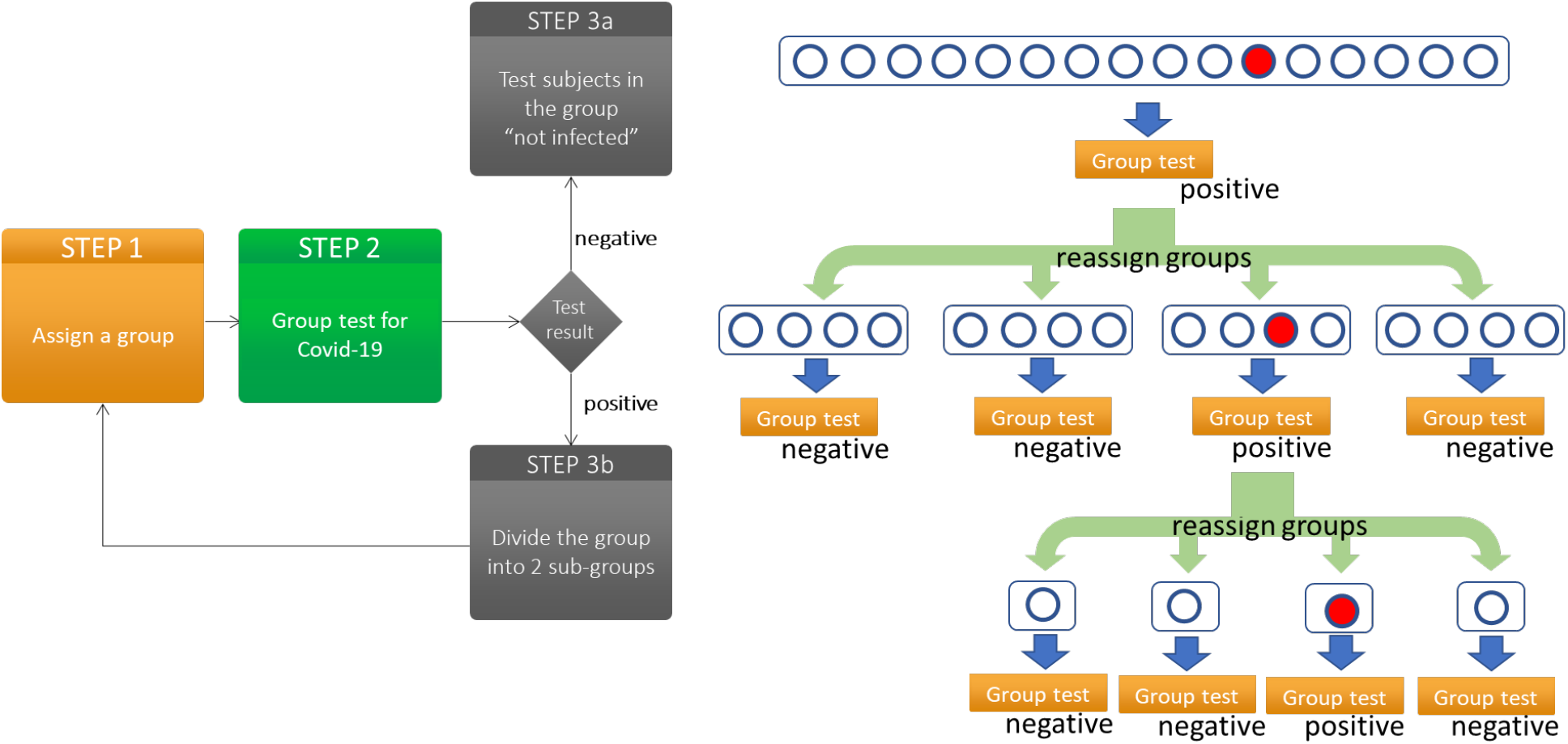
SCALE19 processes and workflow example with the population of 16 with 1 infected test subject

We designed and built an evaluation model that simulates test subject population, infected test subjects according to available COVID-19 statistics, and the group testing processes in SCALE19. The goal of the simulator is to find the required number of tests for different scenarios. Let *P_cv_* be the probability of a patient being confirmed as “infected” for a given population of test subjects. The configurable variables are *N, L, P_cv_*, probability density function (PDF) of infected patients’ locations, geographical characteristics that impact *P_cv_*, and various composition of populations.

## Results

Here, we consider several population models: Generic population, USA population, and the world population. We studied the generic population to understand the impact of several parameters on *T_r_*. We assumed a hypothetical population of N for the generic population model. We then considered a possible reduction of *T_r_* for the actual population of the USA along with its actual reported infection rates. We then considered the world population model with all countries reporting Covid-19 infections.

### Generic Population Model

We first analyzed a generic population model with varying *N, L*, and *P_cv_*. We are interested in *T_r_*, the required number of tests, for the given parameters and how each parameter impacts *T_r_*. We compared this required number of tests with *N* individual tests and find possible savings due to reduction in the number of tests. We ran 100,000 simulated cases with randomly assigned infected patients out of the total population. We first used uniformly distributed infected patients. We also looked at skewed distributions that emulate more realistic cases. We ran 10 batch of simulations to obtain the confidence intervals.

Figure 2 shows *T_r_*, the required number of tests, for varying *P_cv_*. We studied *T_r_* at various *L. N* is fixed at 10,000,000. Different color graphs represent different *L* from 4 to 128.Figure 2.a shows the results when the infected test subjects in the population are distributed uniformly across the population. Thus, different groups would have the same probability of having the infected test subjects. Infected test subjects are evenly distributed among the population and groups would contain probabilistically similar number of infected test subjects. Reduction of *T_r_* is significant when *P_cv_* is less than 0.1, varying from the reduction of 36% to 99%. The positively confirmed cases from tests vary from 0.7% in Vietnam to 30% in United Kingdom according to recent COVID –19 statistics [5]. United Kingdom’s high *P_cv_* is due to its small number of tests and a result of limiting testing to symptomatic subjects. Learning from the statistical trend of other countries, we expect the number of confirmed cases to reduce significantly as the number of tests increases. Also, note that as *L*, group size, increases, the reduction of *T_r_* can increase by as much as 25% for low *P_cv_*.

**Figure 2.**
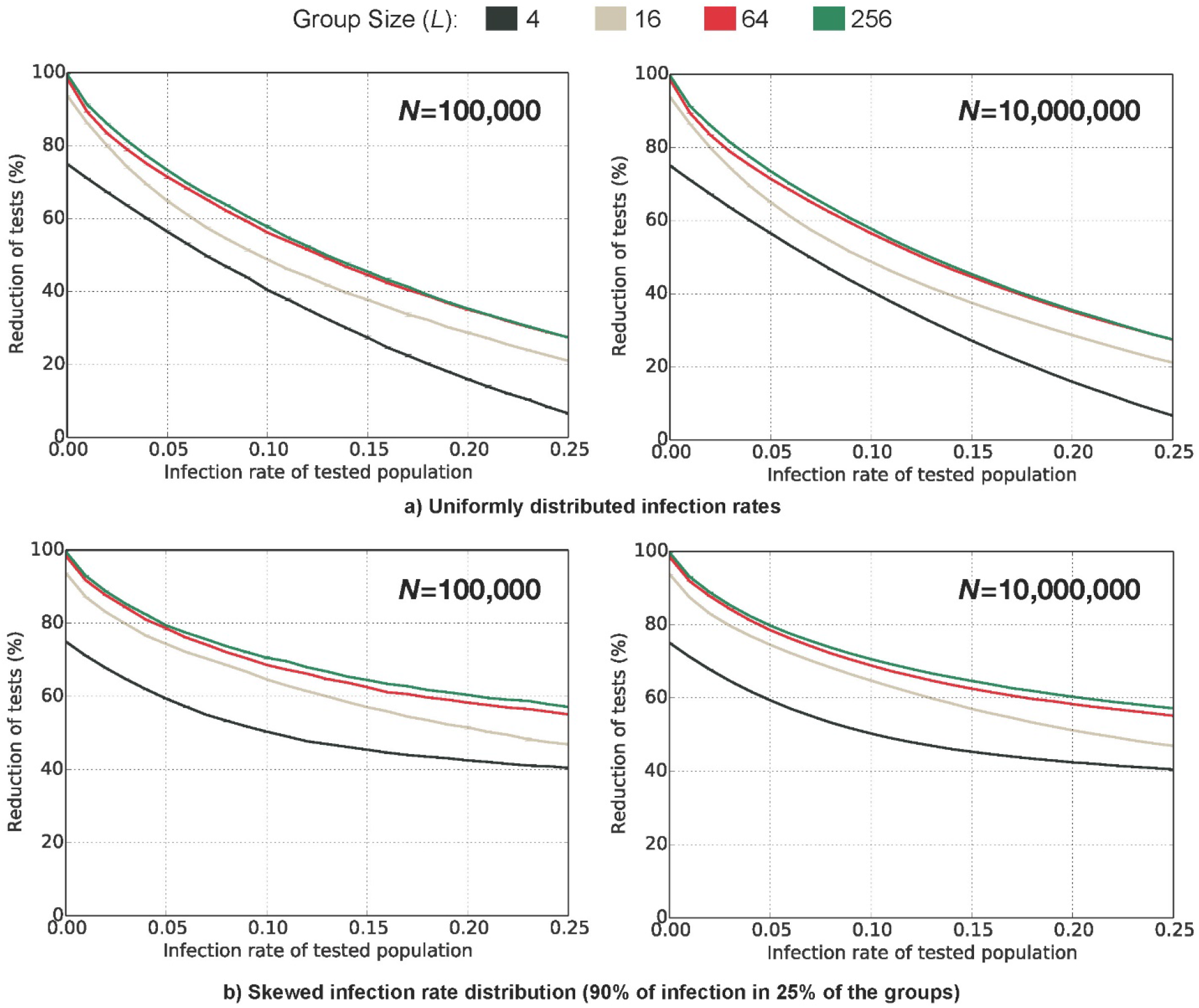
Percentage reduction of the required tests for various L vs. P_i_ for a) uniform infection distribution and b) skewed infection distribution with 25% of population has 90% of infection. Population of 10,000,000.

Figure 2b demonstrates *T_r_* using skewed distribution of the infected population. It represents different geographical characteristics where the infected population is concentrated rather than distributed uniformly. In this figure, we have 25% of the groups containing 90% of infected test subjects. Perhaps such skewed distribution represents the actual population better as the infection tends to cluster around certain geographical regions. Known epicenters are clustered in certain geographical regions, such as New York City, while there are geographical regions with minimal infections. When the infected population is not uniform, *T_r_* is significantly higher for high *P_cv_* than that of the uniformly distributed infection as shown in Figure 2.b. Even with *P_cv_* at 0.25, *T_r_* is reduced by about 30% in the skewed distribution.

Figure 2 shows the impact of *N* (100,000 and 10,000,000) on *T_r_* for uniformly distributed population and skewed distribution. The reduction of *T_r_* does not change much with different *N*, the total population. It shows the savings resulting from SCALE19 can be obtained regardless of the population size.

### USA Population Model

We analyzed SCALE19’s effectiveness using the statistical data obtained from the current cases reported in the USA. We considered each state of the USA separately as their infection rates differ substantially. We divided the population into two categories: tested population and untested population. The tested population corresponds to the total number of people tested up to April 18, 2020 according to Worldometer data [6]. Thus, *N* = 573,223 and *P_cv_* = 233,951 (total confirmed cases)/573,223 (total tests) = 0.4081, for example, for the state of New York. For the untested population, N = 19,618,532 (population of the state of New York) – 573,223 (tested population) = 19,045,309. We need to find *P_cv_* for the untested population. It is difficult to find the right *P_cv_* for the untested population and is an estimation at this point. We analyzed the impact of *P_cv_* on the untested population with several values including a statistic from Iceland. Iceland recently released the results of a population sample of 10,797 Covid-19 tests [7]. These test subjects were invited at random through population screening. They were not part of a high-risk group population (i.e. already symptomatic or returning to Iceland from high-risk countries). 87 test subjects were positive resulting in the value of P_cv_ being 0.8%. We believe 0.8% could possibly represent the infection rate of the untested population. Thus, we used 0.8% infection rate for the untested population.

Figure 3 shows the reduction in *T_r_* for USA with *L* = 16 when 5%, 10%, 20%, 50%, 70%, and 100% of the population are tested. 10% of the population would be 33.1 million. There are currently 3.57 million tested people and we add 29.44 million of the untested population resulting in 10% of the total USA population. For *P_cv_* = 0.8%, the reduction of *T_r_* is significant. Reduction of up to 85% is possible. We show the results for all states separately. As expected, the reduction is lower for the epicenter, New York, where there is a high concentration of infected patients with high *P_cv_*. For USA, up to 280 million tests can be reduced from the total population of 331 million and it would be the equivalent of saving $28 billion, assuming a cost of $100 per test. Figure 3 also shows that the reduction of *T_r_* varies by states. It is certainly higher in the state of New York compared to other states. For example, in the state of Alaska, *T_r_* is only 10,339 tests when 10% of the population is tested.

**Figure 3.**
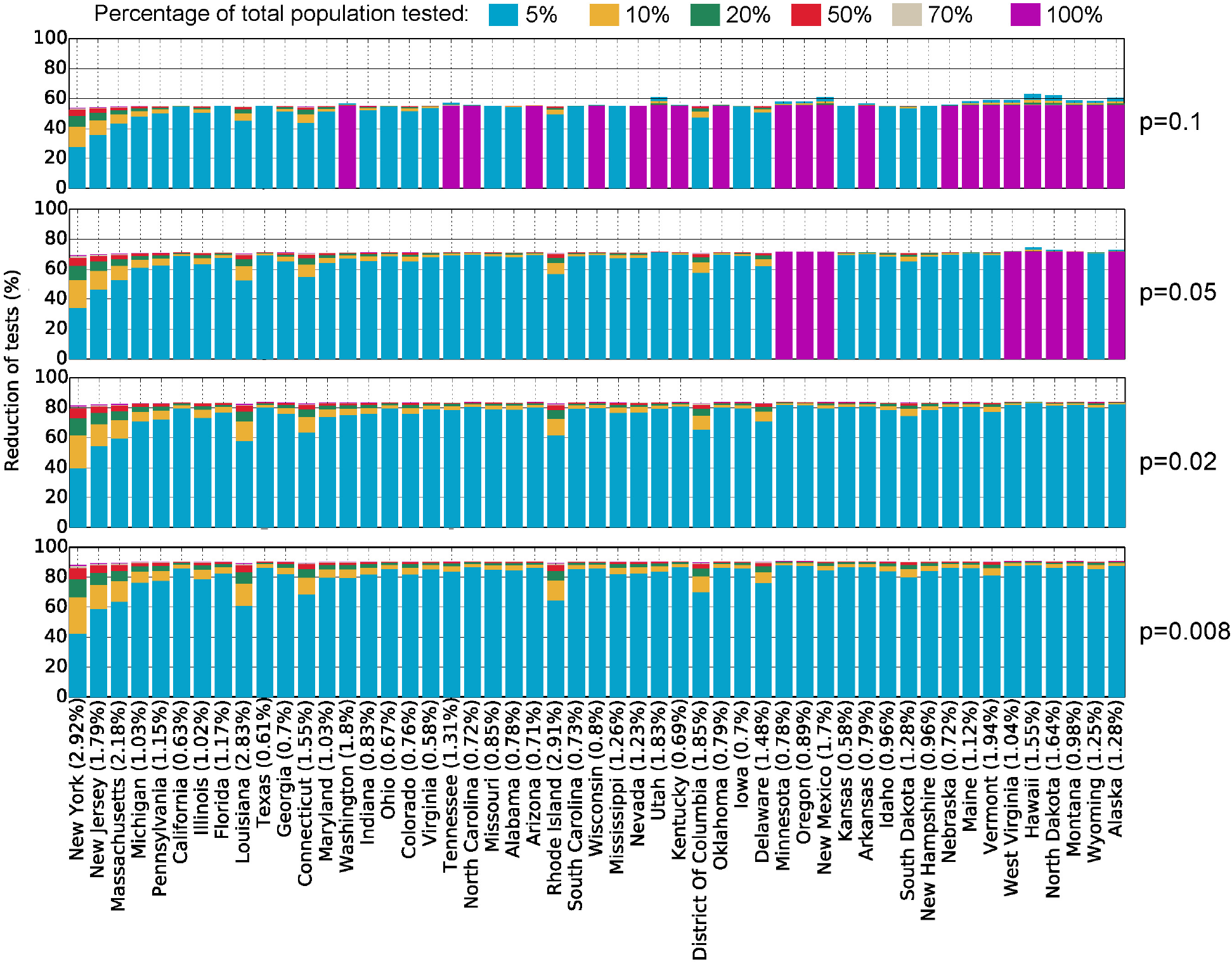
USA Population model: Different colors indicate the reduction in *T*_r_ when 5%, 10%, 20%, 50%, 70%, and 100% of the population are tested.

We also show results for 2%, 5%, and 10% infection rates. Even with *P_cv_* = 0.1, the reduction of *T_r_* is about 60%. When *P_cv_* = 0.02, the reduction of *T_r_* is about 82%, similar to the reduction observed when *P_cv_* = 0.008.

### World Population Model

We analyzed SCALE19’s effectiveness for the world population model. We adopted the same two population models: tested and untested population models for each country. We again assumed 0.8% infection rate for the untested population. Figure 4 shows the reduction in *Tr* for the world with *L* = 16 when 5%, 10%, 20%, 50%, 70%, and 100% of the population are tested. 10% of the world population would be 0.78 billion. There are currently 19.1 million known tested individuals and we add 760.9 million untested people, resulting in 10% of the total world population. With *P_cv_* equal to 0.8% for the untested population, the reduction of *T_r_* is similarly significant.

**Figure 4.**
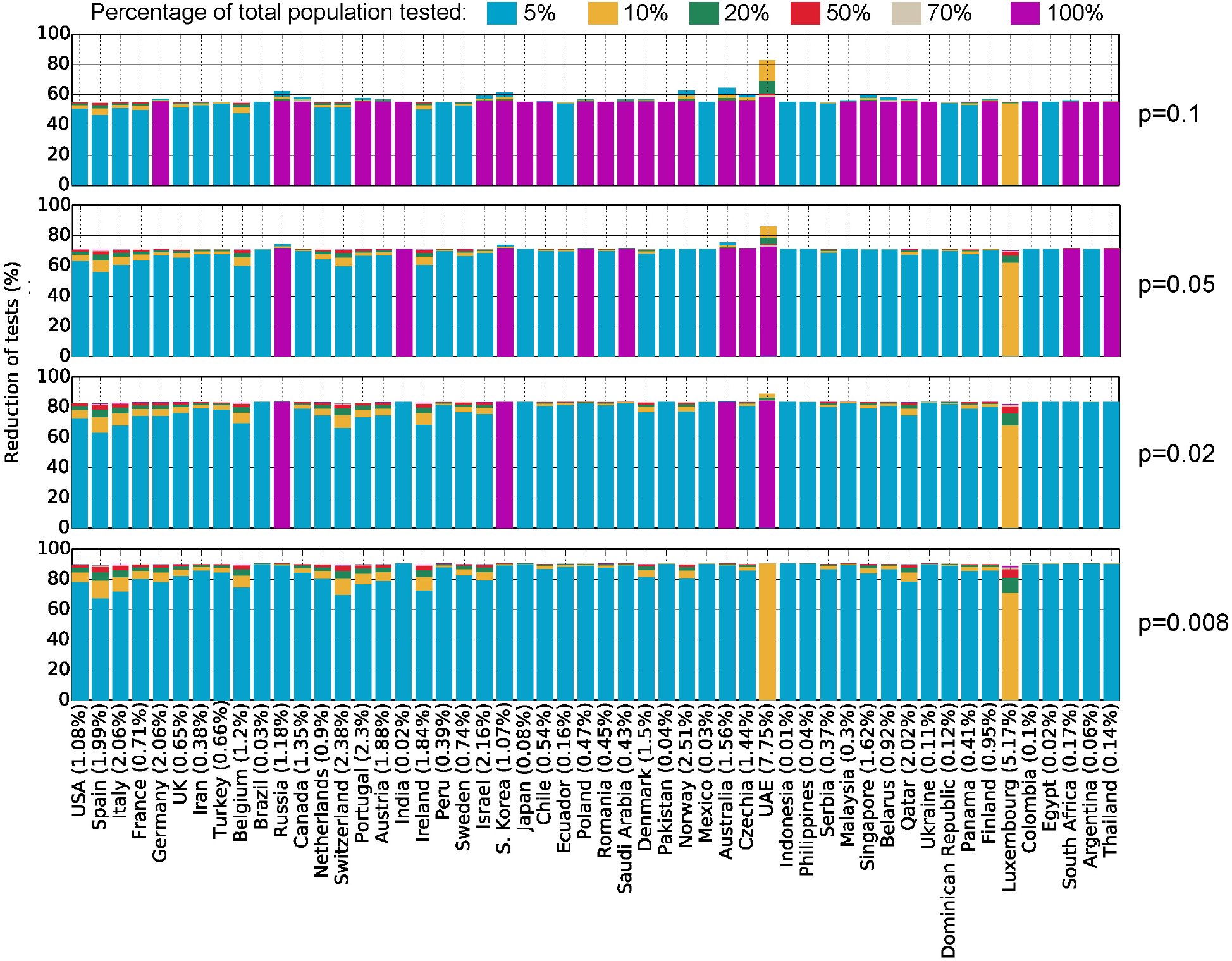
World Population model: different colors indicate the reduction in *T*_r_ when 5%, 10%, 20%, 50%, 70%, and 100% of the population are tested.

Figure 4 shows the top 52 countries in terms of the number of confirmed cases. The rest of the countries are not shown to simplify the graph. It is interesting to find different gains in different countries. This may reflect the varying government policies and each country’s efforts in containing Covid-19. Reduction of up to 89% is possible. For the world, 6.96 billion tests can be reduced from the total population of 7.8 billion and it would be the equivalent of saving $696 billion assuming a cost of $100 per test.

## Discussion

Our study shows that the total required number of tests for global testing could be significantly reduced; the entire population would not have to receive individual testing. For example, in the USA, the total required number of tests can be reduced by 85%, which corresponds to potentially $28 billions in savings. Global testing for the world can be reduced by 89%, which corresponds to potentially $696 billions in savings.

SCALE-19 most significantly reduces number of tests required in populations with low infection rates. Thus, its application may be most efficiently used as a screening tool in lower risk populations. Specific geographical regions, such as rural communities, which may not have been tested before due to availability of testing kits, can now be screened with fewer required tests. More effective and efficient screening for COVID-19 may lead to positive down-stream effects such as reducing usage of personal protective equipment, relaxing of social distancing policies in low risk geographical regions, among others.

We propose that group testing with subsequent recursive testing of sub groups can achieve significant reduction in the number of tests as well as cost savings. However, there are several questions that are not addressed in this study. For numerous known COVID-19 tests, it is reasonable to assume that L = 2,4 even 8 can be achieved without substantially modifying the test process. First, whether a larger grouping (larger *L)* can be applied to currently known Covid-19 tests is not clear. We also need to consider human errors resulting from more complex testing process compared to an individual test. Second, SCALE19 requires multiple samples from the test subject, up to log_2_ *L* samples. It is not clear whether collecting multiple samples and processing of multiple samples can be easily deployed without significant human errors. Automated robotic manipulation of such test samples could potentially address this concern. Third, we cannot confirm a true value for *P_cv_* for the untested population. We conjecture the *P_cv_* value for the purpose of this study by extrapolating from the reported cases, but the question remains. In this study, we do not consider false positive and false negative resulting from the test. We plan to address the impact of tests with false negative and false positive on the reduction and savings in a future study.

We presented a simple algorithm to achieve group testing to identify the presence or absence of infection for each test subject. However, there are more complex algorithms that can improve the reduction of the required number of tests. For example, we can apply combinatorial or probabilistic inference to further reduce the number of required tests. The more complicated the algorithm, the more human errors will impede the accuracy and effectiveness of the testing method. We believe SCALE19 provides a significant reduction in the number of tests and further improvement in the algorithm may not result in much higher reduction due to possible human errors in complex procedures and operations.

## Data Availability

N/A

